# Community and academic hospitals participating in a randomized trial: Patient characteristics, research capacity and trial metrics

**DOI:** 10.64898/2026.01.05.26343452

**Authors:** Vinayak Lad, Alexandra Binnie, Jennifer Tsang, Rosa Myrna Marticorena, Nicole Zytaruk, Diane Heels Ansdell, Deborah Cook

**Affiliations:** Osler Research Institute of Health Innovation, William Osler Health System, Brampton, Ontario, Canada; Department of Critical Care, William Osler Health System, Brampton, Ontario, Canada; Department of Critical Care, Niagara Health Systems, St. Catharines, Ontario, Canada; Department of Medicine, McMaster University, Hamilton, Ontario, Canada; Department of Health Research Methods, Evidence, and Impact, McMaster University, Hamilton, Ontario, Canada; St. Joseph’s Healthcare Research Institute, Hamilton, Ontario, Canada

**Keywords:** community hospitals, research capacity, clinical trials

## Abstract

**Introduction:** Patient populations in Canadian community and academic hospitals may differ with respect to demographic and clinical characteristics, as well as outcomes. Community hospitals may also differ with respect to research capacity and trial performance. The objectives of this study are to examine differences between community and academic hospitals participating in the REVISE trial with respect to: 1) patient characteristics; 2) research capacity, and 3) trial metrics.

**Methods and analysis:** This is a preplanned secondary analysis of the REVISE trial (Re-Evaluating the Inhibition of Stress Erosions), which enrolled patients in 42 ICUs in Canada. Baseline demographic and descriptive outcomes for community and academic hospital patients will be reported using descriptive statistics and compared using t-tests, Wilcoxon rank-sum tests or Chi^2^ test, as appropriate, with p<0.05 indicating statistical significance. To evaluate the impact of site (community versus academic) on the effect of pantoprazole on the primary efficacy and primary safety outcomes, we will use Cox proportional hazards regression analysis, which will be reported as hazard ratio (HR) and 95% confidence intervals, with p<0.05 indicating statistical significance.

**Results:** Results will be of interest to researchers seeking to improve the generalizability of research findings and to policy makers interested in expanding research capacity in Canadian community hospitals.

**Ethics and dissemination:** The REVISE trial was approved by the research ethics boards of the participating hospitals. The protocol for this study will be reviewed by the Hamilton Integrated Research Ethics Board. Findings will be shared through the Canadian Critical Care Trials Group and will be published through conventional academic channels, including abstracts of critical care meetings and peer-reviewed manuscripts.

## Introduction

Canadian hospitals are categorized as academic (or “teaching”) hospitals and community (or “non-teaching”) hospitals [1,2]. Academic hospitals are fully-affiliated with medical schools and frequently participate in research, whilst community hospitals have variable (and often limited) participation in education and research [5-6]. In addition, while academic hospitals frequently offer specialized clinical services, such as cancer care or complex surgical care, to a large catchment area [1-3], community hospitals typically focus on providing acute care services to their local communities [4].

Previous studies have shown that patient populations in community and academic hospitals differ [7]. Patients in community hospitals are older, more likely to be admitted with medical diagnoses, and of lower socioeconomic status [6,8]. They may also have higher severity of illness and higher mortality [7]. These differences call into question whether findings from research studies conducted exclusively in academic hospitals can be generalized to community hospital patients, and vice versa [7].

In this retrospective observational substudy, we will compare the demographic and clinical characteristics of patients enrolled in the Re-Evaluating the Inhibition of Stress Erosions (REVISE) Trial at community and academic hospitals [9,10]. REVISE was an international, multicentre, randomized, placebo-controlled study to determine, among invasively mechanically ventilated patients, the effect of pantoprazole versus placebo on the primary efficacy outcome of clinically important upper gastrointestinal bleeding, and the primary safety outcome of 90-day mortality. The trial found that pantoprazole significantly reduced both clinically important and patient-important upper gastrointestinal bleeding, with no effect on 90-day mortality [10]. Patients in Canada were enrolled in 42 sites, including 30 academic hospitals and 12 community hospitals.

The objectives of this study are to examine differences between community and academic hospitals participating in the REVISE trial with respect to: 1) patient characteristics; 2) research capacity, and 3) trial metrics.

We hypothesize that the community hospital population will be older and have more comorbidities than the academic hospital population. We further hypothesize that clinical outcomes will be similar between settings, but that mortality will be higher in the community hospital population. We hypothesize that the intervention (pantoprazole vs placebo) will not have different effects on the primary efficacy outcome of clinically important upper gastrointestinal bleeding or the primary safety outcome of 90-day mortality between these settings. Regarding research capacity, we hypothesize that community hospitals will have smaller ICU size, fewer full-time equivalent research staff with fewer years of research experience, and fewer ICU studies. Regarding trial metrics, we hypothesize that community hospitals will have lower enrollment, lower informed consent rates, and lower co-enrollment rates but similar protocol adherence.

## Methods

This protocol describes a preplanned analysis of the REVISE trial database (NCT02462590). All patients enrolled in the REVISE trial at one of the 42 Canadian sites will be included. Hospital site type (academic vs community) will be based on the Canadian Institute for Health Information (CIHI) designation of “teaching” (academic) vs “non-teaching” (community) hospital status.

Data on study site characteristics were collected at the time of study site activation and also through a post-study survey administered to research coordinators.

### Analysis

The sample size for this substudy is fixed based on 3265 participants enrolled in Canada, out of a total of 4821 participants in the REVISE Trial.

Patient and site data will be presented using descriptive statistics with means and standard deviations or medians and interquartile range as appropriate for continuous data, and absolute counts and percentages for categorical data.

Community and academic sites will be compared with respect to patient demographics and clinical variables, as well as with respect to research capacity and trial metrics, while accounting for clustering within sites.

To evaluate the impact of community versus academic hospital on the effect of pantoprazole on the primary efficacy outcome (clinically important upper gastrointestinal bleeding) and the primary safety outcome (90-day mortality, we will use Cox proportional hazards regression analysis. Regression results will be reported as hazard ratios (HR) and 95% confidence intervals (CIs). P-values <0.05 will indicate statistical significance.

We will report any missing data pertaining to the reported variables. We will not adjust for multiplicity. Analyses will be performed using SAS 9.4 (Cary, NC).

### Ethics and Dissemination

Protocol implementation for REVISE and database training aligned with the International Council for Harmonisation Guidelines for Good Clinical Practice and other local regulations. The REVISE trial protocol was registered on clinicaltrials.gov (NCT 03374800) and was approved by Health Canada and all research ethics boards associated with participating centers. Free and informed consent was obtained from research participants. The Hamilton Integrated Research Ethics Board, affiliated with McMaster University, was the research board of record for the REVISE trial.

## Results

Results of this study will provide information on similarities and differences between community and academic hospitals participating in a multicenter stress ulcer prophylaxis trial, including characteristics of enrolled patients, the research capacity of participating sites, and trial performance metrics.

## Discussion

This study builds on investigations and policy work that enhance and promote the scientific contribution of research in Canadian community hospitals.

Strengths of this protocol include the large number of patients, the detailed patient clinical and outcome data, and the pre-specified statistical analysis plan. The broad Canadian representation will enhance the generalizability of the findings to other critical care trials across the country.

Limitations of this study include a lack of information on patient ethnicity and sociodemographic variables, which may differ between community and academic patient populations, as these variables were not collected in the original trial database.

We will share results with the Canadian Critical Care Trials Group. We will publish these findings through conventional academic channels, including abstracts of critical care meetings and peer-reviewed manuscripts. We will also disseminate results at professional fora with research trainees.

## Conclusions

This study will fill an evidence gap by generating information on the similarities and differences between patients enrolled in community and academic hospitals in a recent trial of stress ulcer prophylaxis in mechanically ventilated patients. It will also generate information about research capacity and trial performance at community and academic sites participating in a multi-centre randomized trial.

## Data Availability

The REVISE dataset will be used for secondary observational studies such as this one, to address additional hypothesis-driven questions (e.g., predictors of gastrointestinal bleeding). Access by REVISE investigators will follow a submitted rationale, analysis plan and approval by the Management Committee.

## Acknowledgements

The REVISE trial was endorsed by the Canadian Critical Care Trials Group (CCCTG) and the Australian and New Zealand Intensive Care Society. This substudy was conducted as a secondary analysis of the REVISE trial, on behalf of the Canadian Critical Care Trials Group. We appreciate all the patients and family members who participated in this trial, and the work of the Research Coordinators and Site Investigators.

## Funding Statement

The original REVISE trial **was** funded by peer-reviewed grants [Canadian Institutes of Health Research 201610PJT-378226-PJT-CEBA-18373; Canadian Institutes of Health Research 202207CL3-492565-CTP-CEBA-19215]. The National Health and Medical Research Council of Australia grant [GNT1124675] funds enrolment in Australia. REVISE was approved by the National Institute for Health Research (NIHR) in the UK as a Portfolio Study [CPMS ID 45782], eligible for support from the NIHR Clinical Research Network. [https://www.nihr.ac.uk/researchers/collaborations-services-and-support-for-your-research/run-your-study/crn-portfolio.htm]. This study received no support from the commercial or private sector.

## Role of the Funders

The original trial funders and sponsors had no role in the conception, design, conduct, oversight, analysis, interpretation, write-up, review or approval of the manuscript, or decision to submit the manuscript for publication.

## Career Award Funding

D Cook holds a Research Chair in Knowledge Translation in Critical Care from the Canadian Institutes for Health Research.

## Declarations

V.L., A.B., J.T. and R.M.M. are affiliated with Canadian community hospital research programs. The authors declare no additional conflicts of interest related to this work. All authors are affiliated with the REVISE Trial in some capacity.

## Ethics approval and consent to participate

REVISE is approved by Health Canada [HC6-24-c210404], Clinical Trials Ontario Research Ethics Project ID: 1360, the Northern Sydney Local Health District Human Research Ethics Committee (HREC) [2019/ETH08405], and Comissão Nacional de Ética em Pesquisa (CONEP) [5.734.590]. Regulatory oversight is by Health Canada [HC6-24-c210404]. All participating centers had local ethics approval. The Hamilton Integrated Research Ethics Board, affiliated with McMaster University, was the research board of record for the REVISE trial.

## Data Sharing

The REVISE dataset will be used for secondary observational studies such as this one, to address additional hypothesis-driven questions (e.g., predictors of gastrointestinal bleeding). Access by REVISE investigators will follow a submitted rationale, analysis plan and approval by the Management Committee. Requests for access to the dataset by external investigators will be considered following the submission of a rationale, an analysis plan and approval by the Management Committee and research ethics boards, as relevant. Requirements will be stipulated in a pre-specified data sharing agreement.

## Notes

### Competing Interest Statement

The authors V.L., A.B., J.T. and R.M.M. are affiliated with Canadian community hospital research programs.

### Clinical Trial

NCT03374800

### Author Declarations

Hamilton Integrated Research Ethics Board gave ethical approval for the REVISE study.

